# Genetic risk prediction of COVID-19 susceptibility and severity in the Indian population

**DOI:** 10.1101/2021.04.13.21255447

**Authors:** P. Prakrithi, Priya Lakra, The Indian Genome Variation Consortium, Durai Sundar, Manav Kapoor, Mitali Mukerji, Ishaan Gupta

**Author notes:** **Corresponding author** Correspondence to Ishaan Gupta.

## Abstract

Host genetic variants can determine the susceptibility to COVID-19 infection and severity as noted in a recent Genome-wide Association Study (GWAS) by Pairo-Castineira et al.^1^. Given the prominent genetic differences in Indian sub-populations as well as differential prevalence of COVID-19, here, we deploy the previous study and compute genetic risk scores in different Indian sub-populations that may predict the severity of COVID-19 outcomes in them. We computed polygenic risk scores (PRSs) in different Indian sub-populations with the top 100 single-nucleotide polymorphisms (SNPs) with a *p*-value cutoff of 10^−6^ derived from the previous GWAS summary statistics^1^. We selected SNPs overlapping with the Indian Genome Variation Consortium (IGVC) and with similar frequencies in the Indian population. For each population, median PRS was calculated, and a correlation analysis was performed to test the association of these genetic risk scores with COVID-19 mortality. We found a varying distribution of PRS in Indian sub-populations. Correlation analysis indicates a positive linear association between PRS and COVID-19 deaths. This was not observed with non-risk alleles in Indian sub-populations. Our analyses suggest that Indian sub-populations differ with respect to the genetic risk for developing COVID-19 mediated critical illness. Combining PRSs with other observed risk-factors in a Bayesian framework can provide a better prediction model for ascertaining high COVID-19 risk groups. This has a potential utility in the design of more effective vaccine disbursal schemes.

## Introduction

Susceptibility to immune-mediated diseases and viral infections are both observed to be heritable traits, and are associated with specific genetic variants, such as rs11385942, rs10735079^1–5^. The GWAS by Pairo-Castineira et al.^1^ in critically ill COVID-19 patients from a UK cohort identified strong genetic signals, related to antiviral defence mechanisms and inflammatory organ damage, that are potentially associated with COVID-19 severity. Among the top eight robust associations identified in the GWAS^1^, two SNPs, namely, rs10735079 and rs2109069 are also present in the Indian Genome Variation Consortium (IGVC). The IGVC was a large-scale comprehensive study of the Indian sub-populations that was conducted to shed light on the genetic diversity among geographically and ethnically diverse Indian sub-populations^6,7^. This study identified a high degree of genetic distinctness, with respect to SNPs, in different Indian sub-populations^6,7^. With the increasing number of COVID-19 cases in India, a populous and a genetically diverse country, prioritizing vulnerable populations for COVID-19 vaccination is critical, given the limited production of vaccines and identification of genetic risk estimates associated with COVID-19 susceptibility can be beneficial in identifying susceptible population(s).

GWAS have identified the genetic underpinnings of several diseases, and these variants together weighted by their effect sizes yield estimates for polygenic risk score (PRS). PRS provides an estimate of the genetic propensity of an individual to develop a disease and/or a trait^8,9^. Transethnic replication of GWAS effect sizes has been employed previously, however, it is challenging and might not lead to accurate predictions when applied to non-discovery GWAS populations, owing to biological differences, such as different patterns of linkage disequilibrium (LD), allele frequencies, and gene-environment interactions, in different populations^10^ and/or technical differences. For example, there will be no transethnic replication if there is significant difference in the LD structure across different ethnic populations^11^. However, it has been shown that using training data sets which include samples from the discovery population in which the GWAS was conducted (confers the advantage of large sample size in the GWAS) as well as samples from the target population in which the PRS is aimed to be calculated (advantage of being the same ancestry), improves the prediction accuracy of the PRS^12,13^. Hence using the causal variants identified in a discovery GWAS that overlap with the target population instead of taking the SNPs in LD with them, and picking the causal variants having a similar LD pattern across the discovery and target populations, would improve the accuracy of PRS calculated in the target population using the effect sizes of corresponding SNPs from the discovery GWAS.

Here, with the data of stratified Indian sub-populations in hand, we calculated the PRSs with an aim to anticipate Indian sub-populations that are at a higher risk for COVID-19-mediated mortality. Considering the challenges associated with the transferability of the effect sizes, we also analyzed the differences in the patterns of LD, and used the SNPs with similar LD patterns in the discovery population and Indian population to ensure good prediction accuracy of the PRS. Based on these PRSs, we evaluated the population-wise susceptibility that can be of potential utility in more effective vaccine distribution schemes among Indian sub-populations.

## Results

The polygenic predictors used in the present study were derived from Pairo-Castineira et al^1^, and applied on 25 geographically and ethnically diverse sub-populations of the Indian sub-continent^6,7^. However, the existing differences in the LD patterns between different populations can affect the use of the GWAS results in populations different from that in which the genome wide study was performed. Since here we are using the associations derived from a majority European individuals and a few South Asian, East Asian and African individuals to study the risk in the Indian populations, the effect sizes from the GWAS would be ideal to use if the LD pattern around the SNPs used are conserved between the populations. varLD analysis^14^ was performed between Indian and non-Indian ancestral populations of the 1000Genomes project^15^ to assess LD structure across the discovery GWAS population and the target Indian population (Supplementary methods). Almost all the 100 SNPs lie below the threshold (Supplementary Fig. S1), suggesting that the LD structure was maintained between the populations.

As shown in Fig.1a, b and Supplementary Table S1, we found a varying distribution of PRS in different sub-populations of India (one-way ANOVA, F(24, 365) = 3.072, *P* = 2.95 × 10^−06^). In order to determine the relationship between these PRSs and COVID-19 mortality, a correlation analysis was performed. The analysis indicates a positive linear association (Pearson’s correlation coefficient, R = 0.37, and *p* = 0.089) between PRS and COVID-19 mortality (Fig. 2). Based on the PRS for each district, the susceptible population, i.e., the number of individuals in a population at risk for developing severe illness when infected with the SARS-CoV-2 was evaluated (Supplementary table 1). Previously, it was noted that the individuals belonging to different linguistic lineages such as Indo-European (IE), Dravidian (DR), Austro-asiatic (AA), and Tibeto Burman (TB) in different geographic locations of India exhibit genetic distinctness ^6,7^. Here, our results indicate that these subtle genetic differences can affect their susceptibility to COVID-19 mediated illness.

**Fig. 1.**
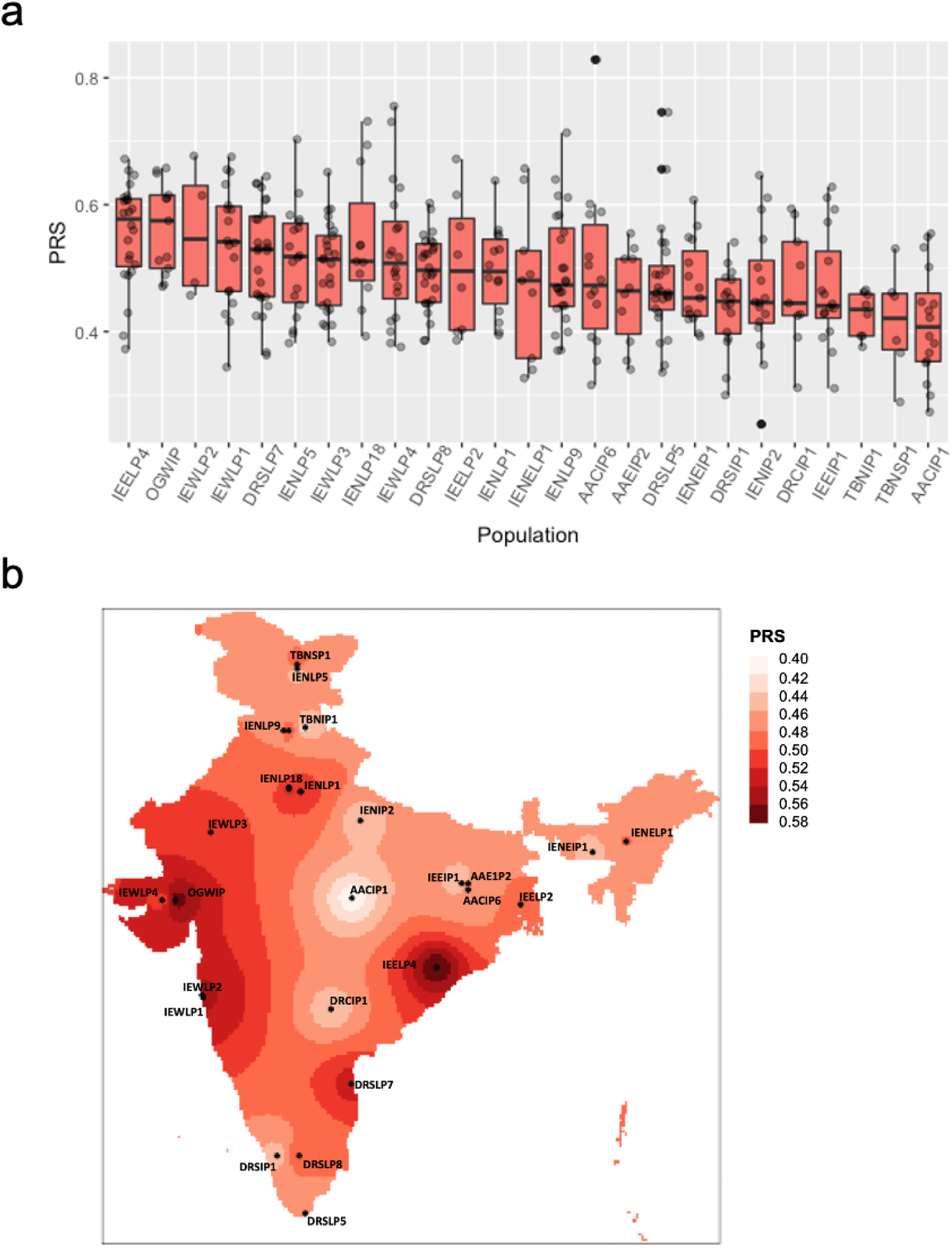
Distribution of polygenic risk score in Indian sub-populations. a) The boxplot is showing polygenic risk score distribution among 25 Indian sub-populations divided on the basis of linguistic and geographical regions (IP, tribal populations; LP, caste; and SP, religious groups). b) Spatial distribution of PRSs for variants in Indian sub-populations spanning different districts of India.

**Fig. 2.**
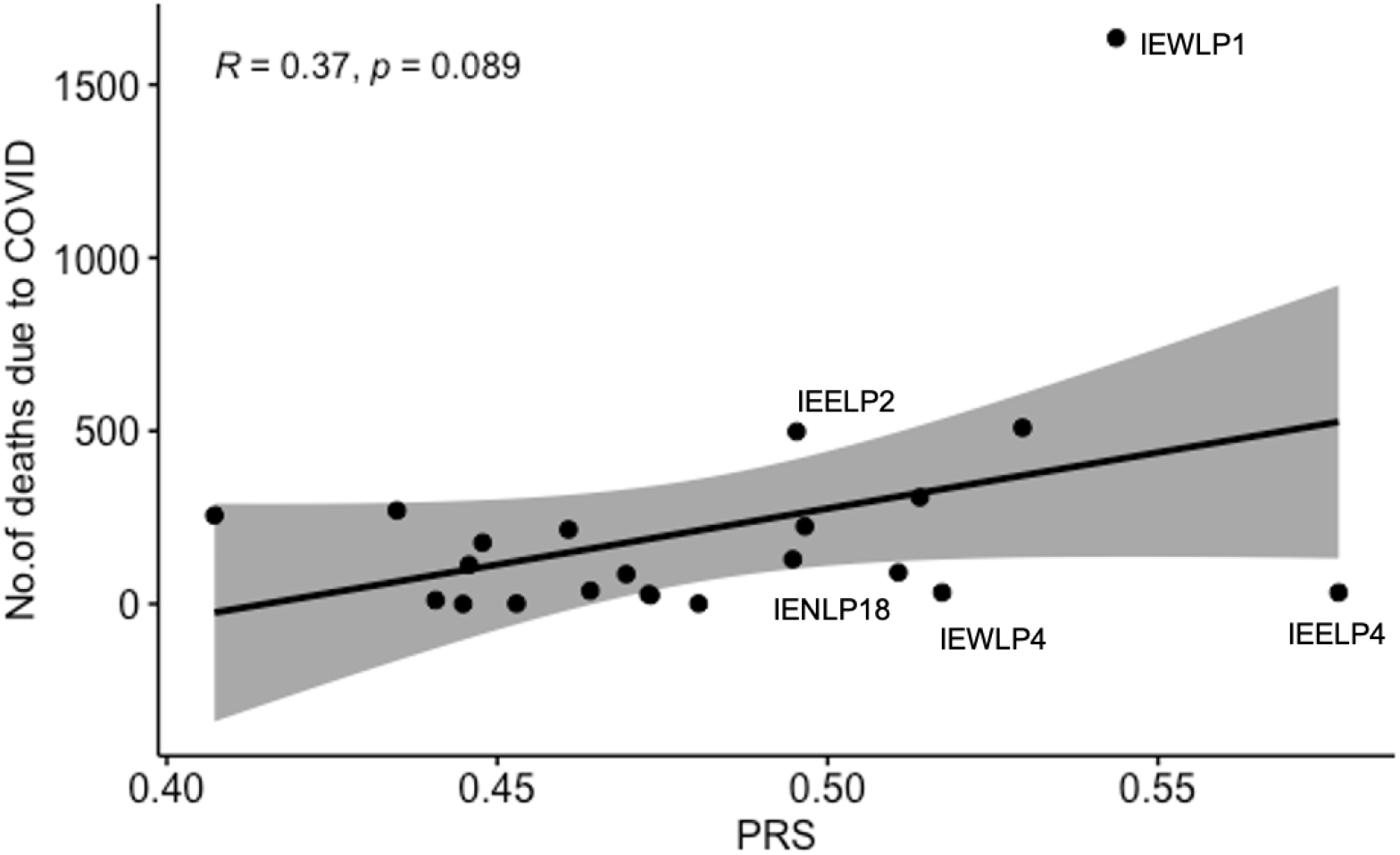
Correlation between COVID-19 mediated deaths and polygenic risk score. Pearson’s correlation analysis showed positive association between PRS and deaths due to COVID-19.

We further investigated whether any non-risk SNP could result in strong correlation with COVID-19 mediated mortality. For this, we selected 100 random SNPs that were not significantly associated with the trait from the same GWAS, and the analyses were repeated multiple times. As shown in supplementary Fig. S2, no strong correlation with the COVID-19 mortality was noted. Our results indicate a correlation of the number of COVID-19-related deaths with the PRS, and thereby can provide support for population specific prioritization in COVID-19 vaccination program. Moreover, our study adds to the existing literature of association between genetics and COVID-19 severity and presents an indication of individuals in Indian sub-populations that are at a high risk of developing critical illness due to COVID-19. These genetic risk scores can, in turn, be employed as a basis of further management of COVID-19 and in the COVID-19 vaccination disbursal scheme.

## Discussion

There are certain limitations of the present study. The GWAS study was not directly conducted in the individuals of the Indian sub-populations, and the PRSs were based on effect sizes from different ancestral groups with COVID-19 infection. The mortality may also be affected by comorbidities like age, diabetes, hypertension, cardiovascular diseases^16–18^ and environmental risk factors that can act as confounders. Further, the current study does not model the effect of confounders such as population density that could also affect the COVID-19 spread and mortality. Smaller populations might have less exposure to the virus, and therefore can display low mortality despite carrying a high PRS. For instance, IEELP4 population (Kandhamal district, population size and density of 7.3 lakhs and 90/km^2^ respectively) has a high PRS but the number of deaths in this district is low, compared to other larger populations (with population size of 10-40 lakhs) with high PRS. Moreover, considering the districts having a similar population size and density (i.e. by removing the outliers), the correlation improved, although not significantly (R=0.43, p=0.092), suggesting that population density could act as a confounding factor. Also, a similar trend was observed in the number of cases over several months in the populations, suggesting that there could be a genetic basis for this trend (Supplementary Fig. S3). The prediction accuracy can be improved by using sequencing data and since IGVC is array data, some of the top causal variants were not represented which could possibly affect the PRS predictions. Increasing the sample size could also provide better accuracy, since IGVC captured only a few individuals of each ancestral group.

In this study, we provide a methodological framework for predicting Indian sub-populations that could be at a higher risk for developing COVID-19 mediated critical illness but not any clinical evidence. These scores in conjunction with the commonly noted comorbidities could provide a good prediction in ascertaining high COVID-19 risk groups. Such accurate identification of vulnerable populations is crucial for the development of effective prevention and vaccination strategies. Such strategies applied to populations with defined genetic histories such as in the Indian subcontinent can be easily extended to model population level susceptibility to several other important diseases that strain the public health system in India, and provide a necessary use case justifying national scale projects such as GenomeIndia.

## Data Availability

For all our analyses, we used the summary statistics available at https://genomicc.org/data. The COVID-19 data for the Indian population was retrieved from ftp://ftp-trace.ncbi.nih.gov/1000genomes/ftp/release/20130502/, https://www.covid19india.org/, https://covid19.Assam.gov.in/district/, https://github.com/covid19india/api. The codes used for the study can be accessed at the following GitHub repository : https://github.com/Prakrithi-P/COVID_PRS_IGV.

https://github.com/Prakrithi-P/COVID_PRS_IGV

## Data availability

For all our analyses, we used the summary statistics available at https://genomicc.org/data,, also provided in the publication of the GWAS used. The COVID-19 data for the Indian population was retrieved from ftp://ftp-trace.ncbi.nih.gov/1000genomes/ftp/release/20130502/, https://www.covid19india.org/, https://covid19.Assam.gov.in/district/, https://github.com/covid19india/api. The codes used for the study can be accessed at the following GitHub repository : https://github.com/Prakrithi-P/COVID_PRS_IGV.

## Funding statement

This work was supported by funds from IITD’s intramural seed grant to IG and in part by a grant from the Department of Biotechnology (DBT), Govt. of India [BT/GenomeIndia/2018] to D.S. and from CSIR [GAP0206] to PP and MM.

## Contributions

IG, MK devised the concept. IG, DS supervised it throughout. PP and IG analysed the data. MM contributed to the IGVC. PL contributed to the figures along with PP. PL, PP, MM and IG wrote the manuscript. All authors read and approved the final manuscript.

## Ethics declarations

### Ethics approval and consent to participate

Not applicable

### Consent for publication

Not applicable

### Competing interests

The authors declare that they have no competing interests.

## Supplementary Information

The Supplementary Information contains Supplementary Figures 1 to 3, Supplementary methods and Supplementary Table 1.

### Rights and permissions

Not applicable

## Supplementary figure and table legends

**Supplementary Fig. S1** Standardized varLD score across CEU and ITU populations. varLD scores for the SNPs analyzed in this study are marked in red, and majority of these are located in the low varLD regions reflecting low differences in LD with respect to these SNPs in these two populations. A similar pattern was observed for the few SNPs whose effect sizes were derived from East-Asian and African ancestral populations with CHB vs ITU and YRI vs ITU respectively.

**Supplementary Fig. S2** Correlation between COVID-19 mediated deaths and polygenic risk score calculated from non-risk SNPs. The *p*-value histogram shows a uniform distribution.

**Supplementary Fig. S3** Number of confirmed cases in different IGV populations over different months. The populations are ordered by those with highest to lowest PRSs.

**Supplementary Table S1** : Population details, PRS and susceptibility data

## Notes

### Competing Interest Statement

The authors have declared no competing interest.

### Funding Statement

This work was supported by funds from intramural seed grant to IG of IITD and in part by a grant from the Department of Biotechnology (DBT) and the Govt. of India - [DBT/GenomeIndia/2018] and a CSIR grant [GAP0206].

### Author Declarations

No patient or participant was involved in the study. Already available genotype data, summary stats and COVID-19 statistics have been applied and used in the bioinformatic analysis.

## References

1. Pairo-Castineira, E. et al. Genetic mechanisms of critical illness in COVID-19. Nature 591, 92–98 (2021).

2. Kenney, A. D. et al. Human Genetic Determinants of Viral Diseases. Annu. Rev. Genet. 51, 241–263 (2017).

3. Kwok, A. J., Mentzer, A. & Knight, J. C. Host genetics and infectious disease: new tools, insights and translational opportunities. Nat. Rev. Genet. 22, 137–153 (2021).

4. Shelton, J. F. et al. Trans-ethnic analysis reveals genetic and non-genetic associations with COVID-19 susceptibility and severity. medRxiv 2020.09.04.20188318 (2020) doi:10.1101/2020.09.04.20188318.

5. Genomewide Association Study of Severe Covid-19 with Respiratory Failure. N. Engl. J. Med. 383, 1522–1534 (2020).

6. Indian Genome Variation Consortium. The Indian Genome Variation database (IGVdb): a project overview. Hum. Genet. 118, 1–11 (2005).

7. Indian Genome Variation Consortium. Genetic landscape of the people of India: a canvas for disease gene exploration. J. Genet. 87, 3–20 (2008).

8. Lewis, C. M. & Vassos, E. Polygenic risk scores: from research tools to clinical instruments. Genome Med. 12, 44 (2020).

9. Chatterjee, N., Shi, J. & García-Closas, M. Developing and evaluating polygenic risk prediction models for stratified disease prevention. Nat. Rev. Genet. 17, 392–406 (2016).

10. Novembre, J. & Barton, N. H. Tread Lightly Interpreting Polygenic Tests of Selection. Genetics 208, 1351–1355 (2018).

11. Martin, A. R. et al. Human Demographic History Impacts Genetic Risk Prediction across Diverse Populations. Am. J. Hum. Genet. 100, 635–649 (2017).

12. Li, Y. R. & Keating, B. J. Trans-ethnic genome-wide association studies: advantages and challenges of mapping in diverse populations. Genome Med. 6, 91 (2014).

13. Márquez-Luna, C., Loh, P.-R. & Price, A. L. Multi-ethnic polygenic risk scores improve risk prediction in diverse populations. Genet. Epidemiol. 41, 811–823 (2017).

14. Ong, R. T.-H. & Teo, Y.-Y. varLD: a program for quantifying variation in linkage disequilibrium patterns between populations. Bioinformatics 26, 1269–1270 (2010).

15. The 1000 Genomes Project Consortium. A global reference for human genetic variation. Nature 526, 68–74 (2015).

16. McGurnaghan, S. J. et al. Risks of and risk factors for COVID-19 disease in people with diabetes: a cohort study of the total population of Scotland. Lancet Diabetes Endocrinol. 9, 82–93 (2021).

17. Guo, W. et al. Diabetes is a risk factor for the progression and prognosis of COVID-19. Diabetes Metab. Res. Rev. e3319 (2020) doi:10.1002/dmrr.3319.

18. Yang, J. et al. Prevalence of comorbidities and its effects in patients infected with SARS-CoV-2: a systematic review and meta-analysis. Int. J. Infect. Dis. IJID Off. Publ. Int. Soc. Infect. Dis. 94, 91–95 (2020).

