## Supplementary figures and methods for "Genetic risk prediction of COVID-19 susceptibility and severity in the Indian population"

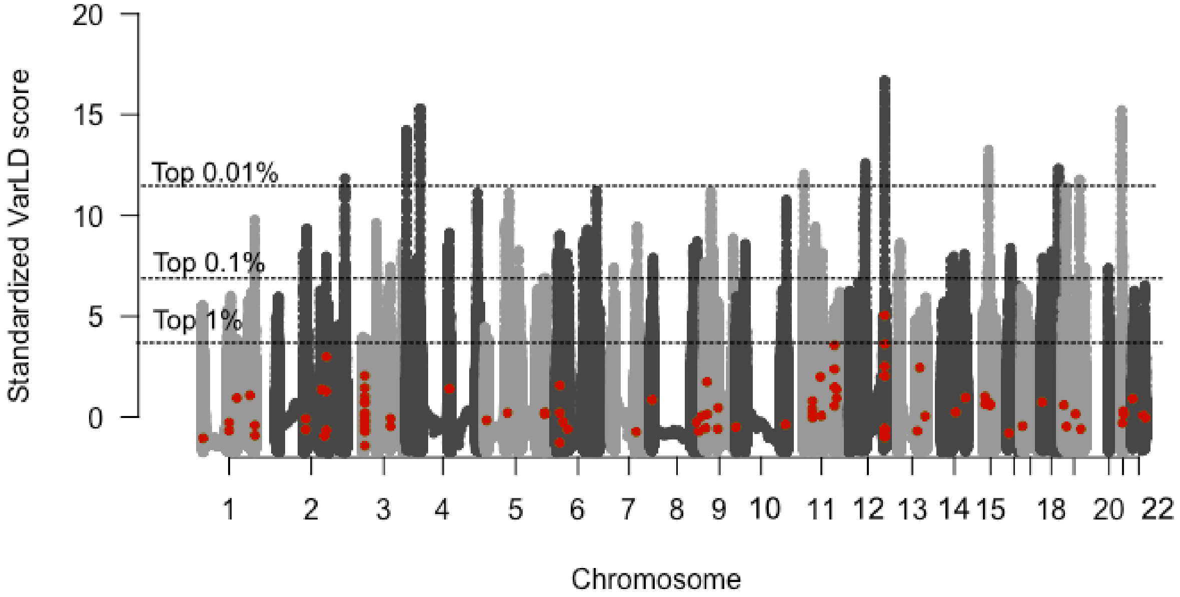


**Supplementary Fig. S1** Standardized varLD score across CEU and ITU populations. varLD scores for the SNPs analyzed in this study are marked in red, and majority of these are located in the low varLD regions reflecting low differences in LD with respect to these SNPs in these two populations. A similar pattern was observed for the few SNPs whose effect sizes were derived from East-Asian and African ancestral populations with CHB vs ITU and YRI vs ITU respectively.

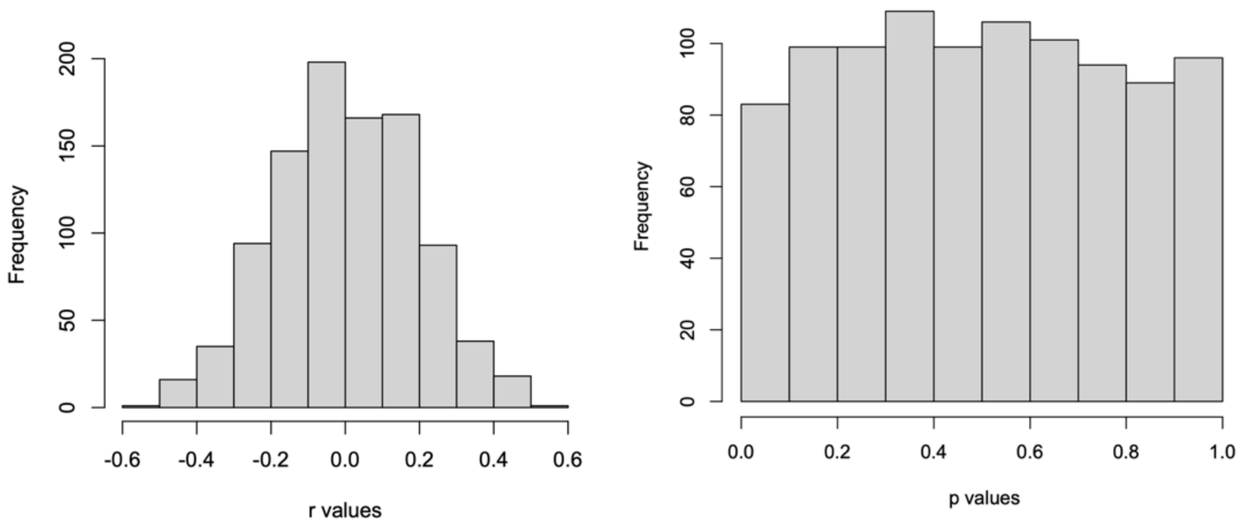


**Supplementary Fig. S2** Correlation between COVID-19 mediated deaths and polygenic risk score calculated from non-risk SNPs. The *p*-value histogram shows a uniform distribution.

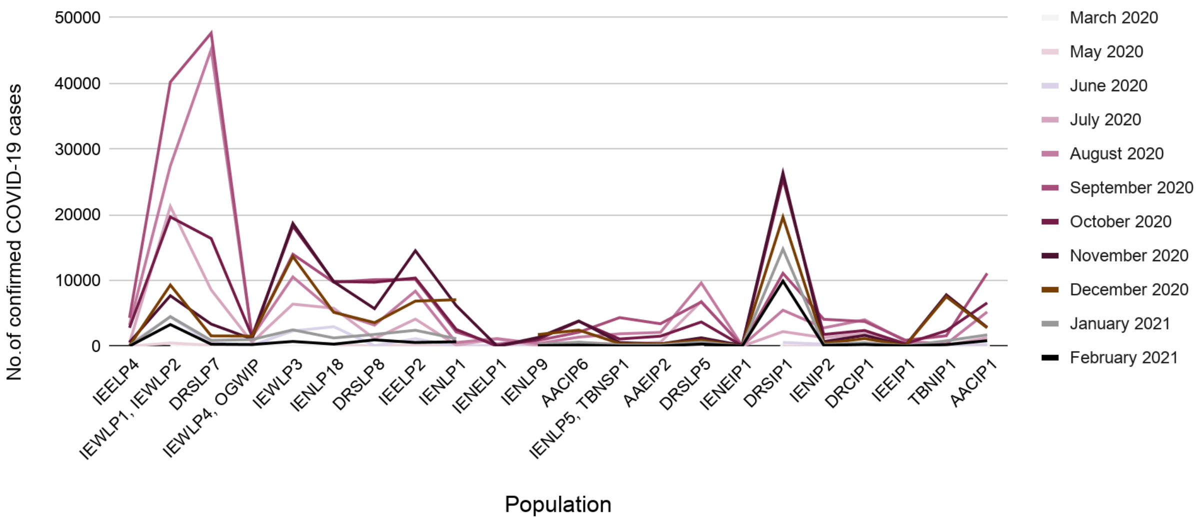


**Supplementary Fig. S3** Number of confirmed cases in different IGV populations over different months. The populations are ordered by those with highest to lowest PRSs.

### Supplementary Methods

#### Study populations and datasets

Summary statistics for genetic variants was obtained from a GWAS study in 2,244 critically ill patients, a majority of them European Ancestry (~75%), ~ 11% of South Asian and 7% of East Asian ancestries. Data of 390 samples from the IGVC was used for analyzing the Indian populations. The detailed description of the populations analyzed and methodology employed in the IGVC is described in their study. Briefly, they represent diverse ethno-linguistic and geographical regions of India, and houses information about genome-wide SNPs across these diverse populations. Here, we have examined 25 different IGVC populations mapping to different districts of India.

#### Linkage disequilibrium

The CEU (individuals from Utah with European ancestry), CHB (Han Chinese in Beijing - representative of European ancestry), YRI (Yoruba in Ibadan, Nigeria - African ancestry) and ITU (Indian Telugu from the UK - representative of the Indian ancestry) populations from the 1000Genomes project (Phase 3) were utilized to compare the patterns of LD (ftp://ftp-trace.ncbi.nih.gov/1000genomes/ftp/release/20130502/). The LD pattern 5MB around the top 100 most significantly associated SNPs were compared between each of these non-Indian ancestral populations with ITU using the varLD (v1.0) tool - a tool to compare the extent of LD differentiation at loci between pairwise populations.

**Polygenic risk score and susceptible population**

The GWAS identified numerous independent genome-wide significant SNPs. These SNPs were overlapped with the IGVC data to identify common variants which were then sorted and filtered on the basis of GWAS *p*-values ($p < 10^{-06}$). The top 100 such SNPs from the study represented in the IGVC data with similar frequencies and LD patterns (for effect sizes from non-SAS ancestry) for the Indian sub-populations were taken for polygenic score analysis. PRS of each individual was calculated using PLINKv1.9, and PRS for a population was calculated by taking median PRS of all the individuals. Population wise statistical significance was calculated using one-way ANOVA. The distribution of the PRS in the individuals across different IGVC populations was plotted using a R script. Total population of each district was multiplied with the corresponding PRS to calculate the potentially susceptible population.

**Correlation analysis**

The district level COVID-19 information till 1st April, 2021 was collected from publicly available repositories including https://www.covid19india.org/, https://covid19.Assam. gov.in/district/, https://api.covid19india.org/. Pearson's correlation between the population median PRS and the number of deaths due to COVID-19 in Indian sub-populations mapped to these districts was calculated using R v4.3.
